# Exome Sequencing for Head and Neck Cancer Predisposition Genes

**DOI:** 10.1101/2025.01.20.25320626

**Authors:** Yao Yu, Bing-Jian Feng, Chun-Pin Esther Chang, Russell Bell, Austin Wood, Ryan Bohlender, Erich M. Sturgis, Guojun Li, Andrew Olshan, Chien-Jen Chen, Pen-Jen Lou, Wan-Lun Hsu, Melissa Cessna, Benjamin Witt, Deb Neklason, Mia Hashibe, Chad D. Huff, Sean V. Tavtigian

**Affiliations:** Department of Epidemiology, Division of Cancer Prevention and Population Sciences, MD Anderson Cancer Center, Houston, TX; Department of Dermatology, University of Utah School of Medicine, Salt Lake City, UT; Division of Public Health, Department of Family and Preventive Medicine, University of Utah School of Medicine, Salt Lake City, UT; Department of Oncological Sciences, University of Utah School of Medicine, Salt Lake City, UT; Department of Otolaryngology - Head & Neck Surgery, Baylor College of Medicine, Houston, TX; Department of Head and Neck Surgery, Division of Surgery, MD Anderson Cancer Center, Houston, TX; Department of Epidemiology, UNC Gillings School of Global Public Health, Chapel Hill, NC; Genomic Research Center, Academia Sinica, Taipei, Taiwan; Department of Otolaryngology, National Taiwan University Hospital, Taipei, Taiwan; Master Program of Big Data in Medical Healthcare Industry, College of Medicine, Fu Jen Catholic University, New Taipei City, Taiwan, and Data Science Center, College of Medicine, Fu Jen Catholic University, New Taipei City, Taiwan; Intermountain Healthcare, Salt Lake City, UT; Department of Pathology, University of Utah School of Medicine, Salt Lake City, UT; Division of Epidemiology, Department of Internal Medicine, University of Utah School of Medicine, Salt Lake City, UT; Department of Epidemiology, UCLA Fielding School of Public Health, Los Angeles, CA

## Abstract

**Introduction:** While genome-wide association studies (GWAS) have identified several common variants associated with head and neck cancer (HNC) risk, large-scale sequence-based studies to evaluate the contribution of rare variants to head and neck cancer risk have not been conducted. The aim of our study was to identify head and neck cancer predisposition genes with exome and targeted sequencing and to assess interactions between the head and neck cancer susceptibility genes and cigarette smoking.

**Methods:** We conducted gene-based and gene-set-based rare-variant case-control analyses to identify germline susceptibility genes for HNC risk. In Phase 1, we generated and analyzed an exome dataset consisting of 220 familial HNC cases to construct a candidate risk gene panel of 501 genes. In Phase 2, we performed targeted sequencing of the candidate gene panel in 2,134 HNC cases and 2,072 controls matched by age, race, and ethnicity. We then conducted rare variant association analysis, incorporating the Phase 2 dataset together with 531 HNC cases and 119,716 cancer-free controls from UK Biobank whole-exome sequencing data via meta-analysis. We then estimated effect sizes of rare variant classes in known and newly implicated HNC susceptibility genes and pathways for HNC risk.

**Results:** We identified 27 genes with nominally significant meta p < 0.05 among the 501 genes evaluated in Phase 2, including six known cancer predisposition genes, *BRCA1*, *BRCA2*, *RAD51B*, *BAP1*, *APC*, and *MUTYH*. Loss-of-function (LoF) variants in *BRCA1* (OR=5.0, 95% CI: 1.72-14.57), *BRCA2* (OR=2.56, 95% CI: 1.22-5.41), and *MUTYH* (OR=4.84, 95% CI: 1.01-13.86) exhibited significantly elevated effect sizes. Individuals who only smoked had an OR for head and neck cancer risk of 2.97 (95%CI=2.65, 3.33), individuals who only carried LoF or predicted damaging variants in the DNA homologous recombination repair genes had an OR of 1.74 (95%CI=1.14, 262), and individuals who were carriers and smokers had an OR of 5.24 (95%CI=3.74, 7.32).

**Conclusions:** We observed associations with HNC risk for DNA repair pathway genes associated with breast cancer and polyposis colorectal cancer. We also observed suggestive interactions between these variants in these genes and cigarette smoking. Our results indicate that smokers with pathogenic variants in the DNA repair genes implicated in this study are at ∼5-fold risk of developing HNC.

## Introduction

Each year, approximately 771,000 head and neck cancer cases are diagnosed throughout the world.^1^ Head and neck cancer includes cancer subsites of the oral cavity, oropharynx, hypopharynx, and larynx. The highest burden in the world of incident cases of head and neck cancer is in India (223,000 cases), China (84,000 cases) and the US (56,000 cases). High age-standardized incidence rates of head and neck cancer are observed in Russia (9.9 per 100,000), the UK (9.8 per 100,000), and the United States (9.8 per 100,000). Tobacco use and alcohol consumption are well-established risk factors, with attributable risks ranging from 51% for North America, to 83% for South America, and 84% for Europe.^2^ HPV infections are a strong risk factor for oropharyngeal cancer.^3^

Previous array-based genome-wide association studies (GWAS) in head and neck cancer have identified independent common genetic variants associated with cancer risk, near genes involved in the alcohol and tobacco metabolism and DNA repair pathways. A GWAS on European head and neck cancer patients (2,091 cases and 3,513 controls) reported associations with variants in the *ADH* gene cluster, near *ALDH2* and near the DNA repair gene *HEL308*.^4^ With additional cases and controls (6,034 cases and 6,585 controls), 8 loci were associated with oral and pharyngeal cancers (6p21.32, 11p15.4), oral cancer (2p23.3, 9q34.12, 9p21.2, 5p15.33) and oropharyngeal cancer (HLA region).^5^ In the Million Veteran Program Cohort, variants associated with HNC risk were identified from a case-control study of 4,012 HNC cases and 16,048 controls: loci in *DMBT1* and *TPM1* were associated with oral cavity, hypopharyngeal and laryngeal cancer while loci in *CRTAM*, *CHD5,* and *CDH2* were associated with oropharyngeal cancer.^6^ In this cohort, variants associated with HNC risk for African-American patients were also identified in immune genes such as *HLA-G*, *GA1*, *CD6*, *NCAM1*/*CD56*, *CD68*, *SOCS6*.^6^ Another GWAS in 2,171 non-Hispanic white head and neck cancer cases and 4,493 controls reported on associations with *HLA-DQB1*, *CCHCR1*, and a novel 6p22.1 locus.^7^ Several GWAS and large-scale genotyping studies have been conducted in Asian and African American populations. A GWAS study on 993 Chinese laryngeal cancer patients and 1,995 controls, with replication in 2,398 cases and 2,804 controls reported on three loci: *FADS1*, *AIF1*, and *TBX5*.^8^ Another GWAS study on 1,529 Taiwanese oral cancer patients and 44,572 controls reported associations with variants in the *HLA-B* and *HLA-DQ* gene cluster, as well as in *TERT-CLMPT1L*, *ADH1B*, *LAMC3*, *SKIV2L* and *TNXB* genes.^9^

In contrast to array-based GWAS, large-scale sequence-based studies of head and neck cancer risk have not previously been conducted, and thus, little is known about the contribution of rare, protein-coding variants to head and neck cancer risk. Our study generated and analyzed exome and targeted sequencing data to identify head and neck cancer predisposition genes with an excess of rare, putatively dysfunctional variants in cases relative to controls. In addition, we assessed potential interactions between the identified predisposition genes and cigarette smoking.

## Results

Phase 1 included 220 familial HNC cases, while Phase 2 was comprised of 2,134 HNC cases and 2,072 controls (Table 1). In both phases, a high proportion of cases were diagnosed with oropharyngeal cancer (39.5% in Phase 1 and 29.8% in Phase 2). More than 70% of cases and controls were males. Phase 2 included a high number of Asian and Pacific Islander cases, reflecting the recruitment of a large proportion of participants from the Taiwan center. As expected, the percentage of individuals who were ever cigarette smokers or alcohol drinkers was higher among cases in both Phase 1 and Phase 2, compared to controls in Phase 2.

**Table 1.** Characteristics of familial head and neck cancer (HNC) cases in Phase 1 and HNC cases and controls in Phase 2 of the study.

|  | Phase 1<br>(N=220) |  | Phase 2<br>(N=4,206) |  |  |  |
| --- | --- | --- | --- | --- | --- | --- |
|  | Familial<br>HNC cases |  | HNC Cases<br>(N=2,134) |  | Controls<br>(N=2,072) |  |
|  | N | % | N | % | N | % |
| **Study center** |  |  |  |  |  |  |
| Houston | 96 | 43.6 | 680 | 31.9 | 781 | 37.7 |
| North Carolina | 63 | 28.6 | 1,053 | 49.3 | 914 | 44.1 |
| Taiwan | 13 | 5.9 | 401 | 18.8 | 377 | 18.2 |
| Utah | 48 | 21.8 | NA |  | NA |  |
| **Cancer subsite** |  |  |  |  |  |  |
| Oral Cavity | 63 | 28.6 | 514 | 24.1 |  |  |
| Oropharynx | 87 | 39.5 | 635 | 29.8 |  |  |
| Hypopharynx | ** |  | 146 | 6.8 |  |  |
| Oral Cavity - Oropharynx-Hypopharynx, NOS | ** |  | 317 | 14.9 |  |  |
| Larynx | 44 | 20.0 | 522 | 24.5 |  |  |
| **Sex** |  |  |  |  |  |  |
| Female | 60 | 27.3 | 447 | 20.9 | 534 | 25.8 |
| Male | 160 | 72.7 | 1,687 | 79.1 | 1,538 | 74.2 |
| **Age (years)** |  |  |  |  |  |  |
| <55 | 72 | 32.8 | 827 | 38.8 | 817 | 39.4 |
| 55 to <65 | 78 | 35.4 | 711 | 33.3 | 649 | 31.3 |
| 65+ | 70 | 31.8 | 596 | 28.0 | 606 | 29.2 |
| **Race/ethnicity** |  |  |  |  |  |  |
| Non-Hispanic White | 175 | 79.5 | 1,421 | 66.6 | 1,414 | 68.2 |
| Black/African American | 24 | 10.9 | 259 | 12.1 | 208 | 10.0 |
| Hispanic | ** |  | 30 | 1.4 | 57 | 2.8 |
| Asian and Pacific Islanders | ** |  | 405 | 19.0 | 382 | 18.4 |
| **Cigarette smoking** |  |  |  |  |  |  |
| Never cigarette smoker | 58 | 26.4 | 441 | 20.7 | 950 | 45.8 |
| Ever cigarette smoker | 152 | 69.1 | 1,690 | 79.2 | 1,122 | 54.2 |
| **Alcohol drinking** |  |  |  |  |  |  |
| Never alcohol drinkers | 47 | 21.4 | 477 | 22.4 | 839 | 40.5 |
| Ever alcohol drinkers | 162 | 73.6 | 1,654 | 77.5 | 1,233 | 59.5 |

\*Other included non-specific or unknown race
\*\* suppressed since one of the cell counts was <11

We conducted whole-exome sequencing on Phase 1 cases with a targeted depth of 60X for each sample. We then performed two analyses on these cases to identify HNC susceptibility genes (Figure 1). First, we conducted joint genotype calling with samples from the 1000 Genomes Project (G1K).^10^ We then performed gene-based and gene-set-based burden tests using VICTOR^11^ followed by a gene prioritization analysis that combined gene-based burden test p-values with biologically relevant scores.^12^ Second, we conducted an extended case-control analysis with additional cases from the Cancer Genome Atlas Head-Neck Squamous Cell Carcinoma (TCGA-HNSC) data, cancer-free controls sequenced at the University of Texas MD Anderson Cancer Center, and unaffected parents from the National Database for Autism Research (NDAR). After joint genotype calling following GATK’s best practice, we conducted cross-platform quality control using XPAT.^13^ We then conducted gene-based and gene-set-based association tests using VAAST2^14^ and PheVor^15^ to combine biomedical ontologies to identify HNC-associated genes. We consolidated the top 100 genes from each analysis, together with a list of known or suspected cancer susceptibility genes, into a panel of candidate genes. As a result, we obtained a list of 501 candidate genes for HNC risk and designed a targeted sequencing panel.

**Figure 1.**
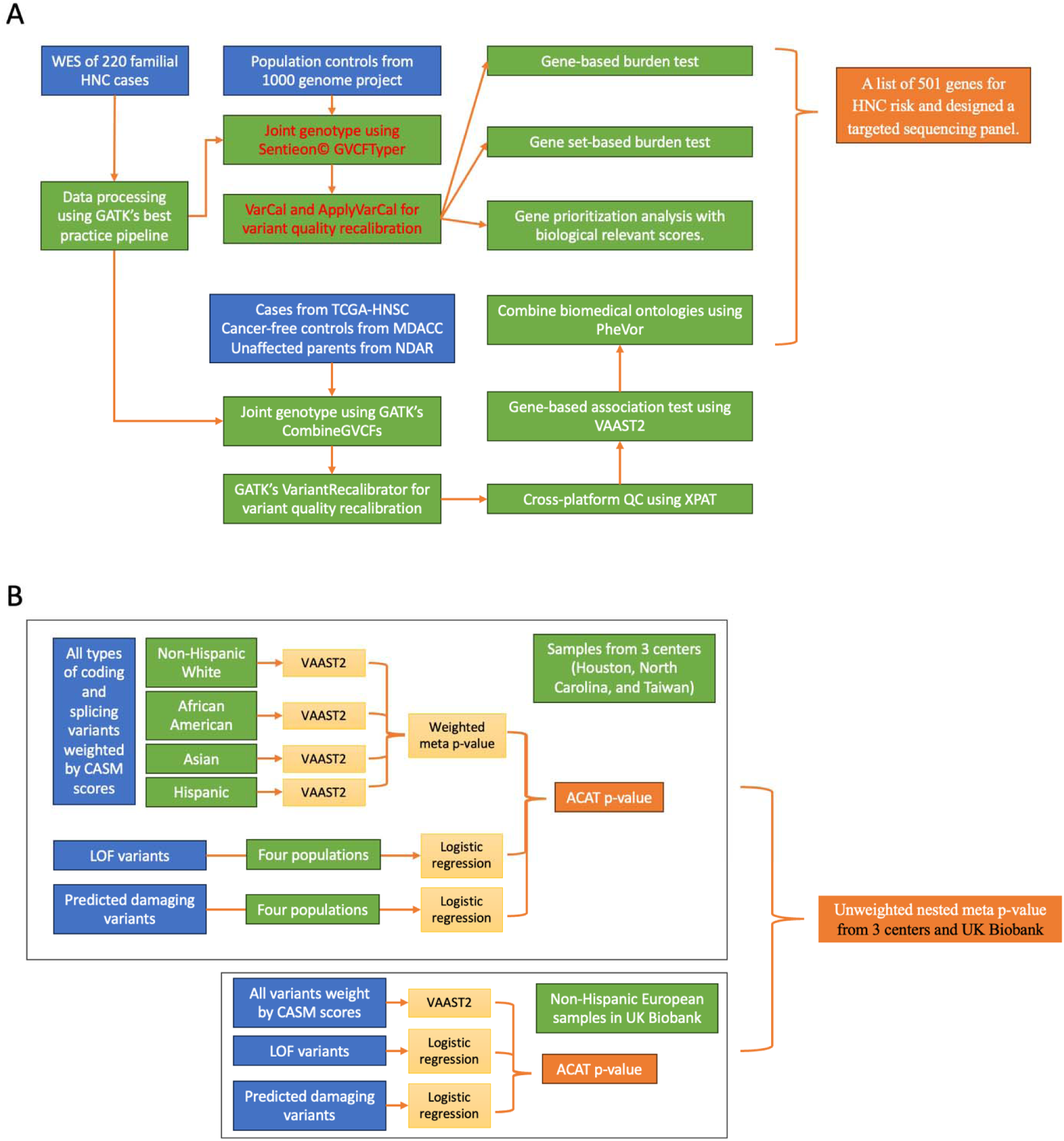
Overview of the gene-based association study design using targeted sequencing data from three centers and whole-genome sequencing data from UK Biobank. (A) Study design for Phase 1 analysis. (B) Study design for Phase 2 analysis.

We then performed targeted sequencing on 2,134 HNC cases and 2,072 controls recruited in Phase 2, followed by a gene-based association analysis. Phase 2 HNC cases and controls were recruited from the study centers in Houston, North Carolina, and Taiwan (3-center data). To ensure the quality of the genotypes, we conducted joint genotype calling using GATK’s best practice pipeline^16^ with VariantRecalibrator for variant quality score recalibration and using Sentieon© GVCFTyper with VarCal and ApplyVarCal for variant quality recalibration. We compared the results from the above two pipelines and included only the consistent genotypes in the subsequent analysis. We then implemented rigorous quality control measures to ensure high data quality, retaining only consistent and high-confidence genotypes for analysis. This process identified 175,277 single-nucleotide variants (SNVs) and 5,518 insertions/deletions (INDELs) with minor allele frequencies (MAF) < 0.005. Among these, 34,308 variants were located in protein-coding or splicing regions. Among the exonic variants, 1,004 were loss-of-function (LoF) variants (including nonsense, frameshift INDELs, and splicing-altering variants), 373 were in-frame INDELs, and 19,887 were missense variants.

Next, we conducted gene-based association tests on the 461 genes out of the 501 ones included in Phase 2 targeted sequencing panel (requiring each with at least four scorable variants) across four populations: non-Hispanic whites (NHW), African Americans (AA), Hispanics (HIS), and Asians (ASI). We conducted rare variant association tests with all variants, LoF variants, and predicted damaging variants, followed by an aggregated Cauchy association test (ACAT) to integrate these tests (see details in Figure 1 and Methods). To account for analyses across the four populations, we conducted a meta-analysis, combining ACAT p-values and weighting them by the effective sample size of each population. In the multi-population meta-analysis, we observed nominally significant associations in several known cancer predisposition genes, including *RAD51B* (p = 0.006), *BAP1* (p = 0.0223), and *BRCA1* (p = 0.0462) (Table 2 and Supplementary Table S1).

**Table 2.**
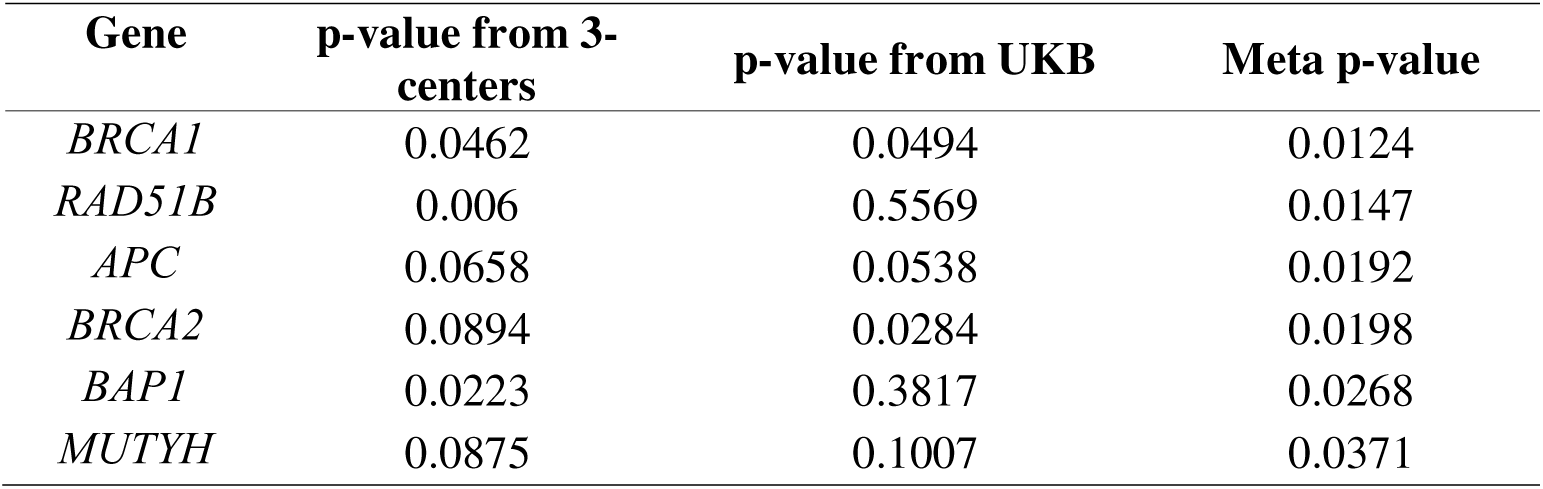
P-values for association with head and neck cancer risk for known cancer predisposition genes combining results from the 3-center data (Houston, North Carolina, and Taiwan) and UK Biobank.

| **Gene** | **p-value from 3-**<br>**centers** | **p-value from UKB** | **Meta p-value** |
| --- | --- | --- | --- |
| *BRCA1* | 0.0462 | 0.0494 | 0.0124 |
| *RAD51B* | 0.006 | 0.5569 | 0.0147 |
| *APC* | 0.0658 | 0.0538 | 0.0192 |
| *BRCA2* | 0.0894 | 0.0284 | 0.0198 |
| *BAP1* | 0.0223 | 0.3817 | 0.0268 |
| *MUTYH* | 0.0875 | 0.1007 | 0.0371 |

To validate our findings using targeted gene sequencing data from Phase 2 samples, we extended the association analysis to include HNC cases and cancer-free controls from the UK Biobank.^17^ We obtained whole-exome sequencing data from 531 NHW cases and 119,716 NHW cancer-free controls and performed joint genotype calling using DeepVariant implemented in GLnexus,^18^ followed by quality control using XPAT. Matching the approach described above, we applied the omnibus test exome-wide, incorporating analyses of all coding variants with prioritization scores from VAAST2, LoF variants, and LoF with predicted damaging variants. Two of the six known cancer predisposition genes identified in Phase 2 were nominally significant: *BRCA1* (p = 0.0494) and *BRCA2* (p = 0.0284).

We then conducted a meta-analysis using the 3-center data and UK Biobank data for the targeted genes. We observed no inflation in the p-values in the meta-analysis (Supplementary Fig. S1). The six known cancer predisposition genes previously identified in Phase 2 are nominally significantly associated with HNC risk (p < 0.05), including *BRCA1* (p = 0.0124), *BRCA2* (p=0.0198), *RAD51B* (p = 0.0147), *BAP1* (p = 0.0268), *APC* (p=0.0192), and *MUTYH* (p=0.0371). Additionally, we identified 16 novel candidate HNC risk genes with meta p-value < 0.05 (Table 3). Using UK Biobank data exclusively, we also observed several genes associated with HNC risk under a suggestive significance level of 0.001, which were not part of the targeted sequencing panel, including *PCDHB5*, *TANC1*, and *ADGRA2* (Table 4).

**Table 3.**
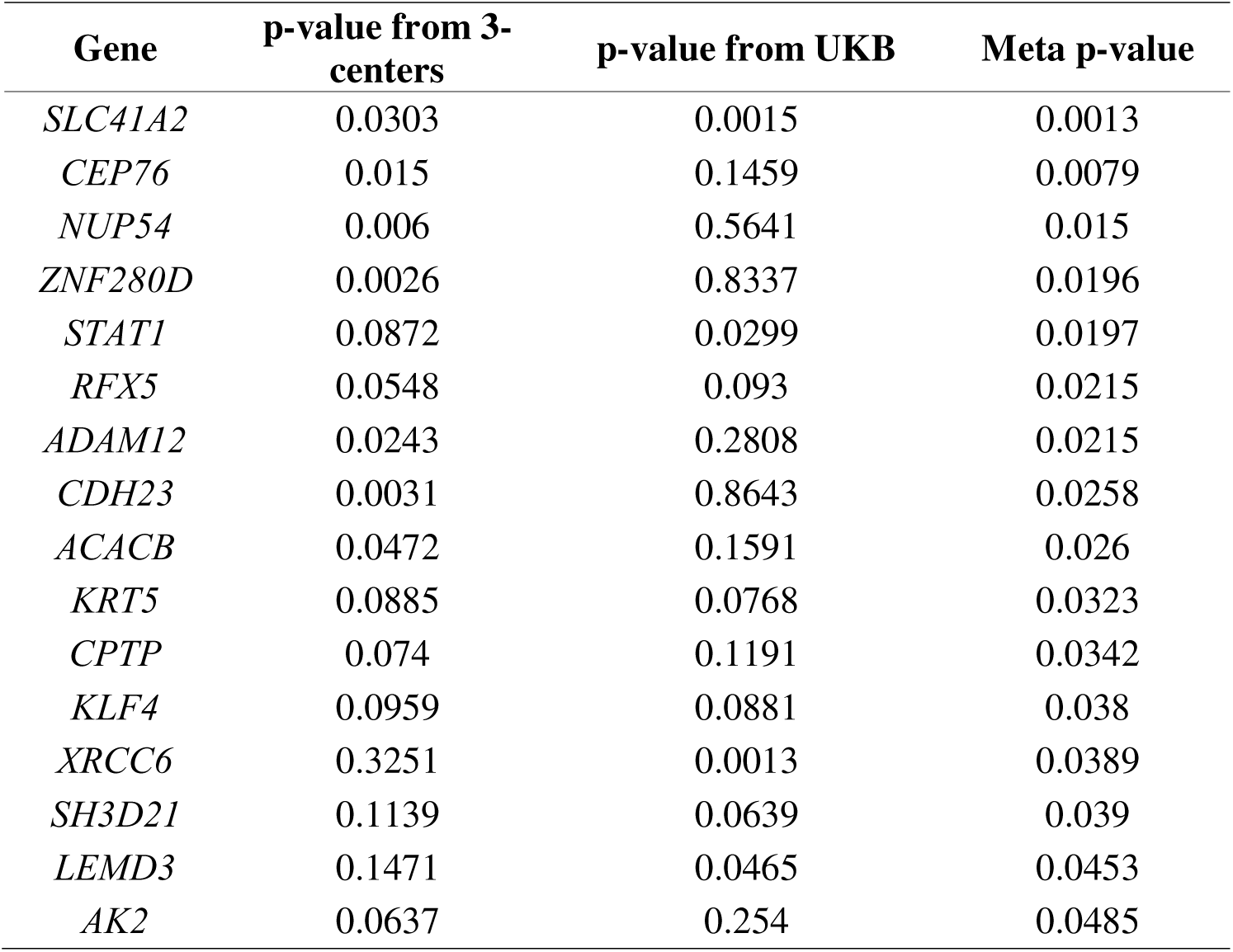
P-values for association with head and neck cancer risk for genes with Meta p-value < 0.05 combining results from the 3-center data (Houston, North Carolina, and Taiwan) and UK Biobank.

| **Gene** | **p-value from 3-centers** | **p-value from UKB** | **Meta p-value** |
| --- | --- | --- | --- |
| *SLC41A2* | 0.0303 | 0.0015 | 0.0013 |
| *CEP76* | 0.015 | 0.1459 | 0.0079 |
| *NUP54* | 0.006 | 0.5641 | 0.015 |
| *ZNF280D* | 0.0026 | 0.8337 | 0.0196 |
| *STAT1* | 0.0872 | 0.0299 | 0.0197 |
| *RFX5* | 0.0548 | 0.093 | 0.0215 |
| *ADAM12* | 0.0243 | 0.2808 | 0.0215 |
| *CDH23* | 0.0031 | 0.8643 | 0.0258 |
| *ACACB* | 0.0472 | 0.1591 | 0.026 |
| *KRT5* | 0.0885 | 0.0768 | 0.0323 |
| *CPTP* | 0.074 | 0.1191 | 0.0342 |
| *KLF4* | 0.0959 | 0.0881 | 0.038 |
| *XRCC6* | 0.3251 | 0.0013 | 0.0389 |
| *SH3D21* | 0.1139 | 0.0639 | 0.039 |
| *LEMD3* | 0.1471 | 0.0465 | 0.0453 |
| *AK2* | 0.0637 | 0.254 | 0.0485 |

**Table 4.** P-values for association with head and neck cancer risk for genes not included in the Phase 2 targeted sequencing panel with p<0.001 from the UK Biobank.

| Gene | p from UKB* |
| --- | --- |
| *PCDHB5* | 9.93E-06 |
| *TANC1* | 4.64E-05 |
| *ADGRA2* | 6.58E-05 |
| *KDM8* | 0.0002 |
| *LACTBL1* | 0.0002 |
| *B4GALNT2* | 0.0002 |
| *AGER* | 0.0003 |
| *CDC6* | 0.0007 |
| *FZD7* | 0.0007 |
| *NPAT* | 0.0008 |
| *CHTOP* | 0.0008 |
| *CDR2* | 0.0008 |
| *SLPI* | 0.0008 |

\*Threshold for the suggestive significant p-value was calculated as $0.05/(19500/500)$

To characterize the contribution of rare genetic variation to the risk of HNC, we evaluated the effect sizes of LoF variants and predicted damaging variants in NHW (Figure 2 and Supplementary Fig. S2-S5). The LoF variants in *BRCA1* (OR=5.0, 95% CI: 1.72-14.57), *BRCA2* (OR=2.56, 95% CI: 1.22-5.41), and *MUTYH* (OR=4.84, 95% CI: 1.01-13.86) exhibited significantly elevated effect sizes. The LoF variants in *APC*, *BAP1*, and *RAD51B* also exhibited moderate-to-large odds ratios (5.36, 11.95, and 2.11, respectively), though with wide confidence intervals due to sample sizes and variant rarity. In contrast, the odds ratios for missense variants in these genes were only modestly elevated and were not statistically significant. LoF variants in other top genes from UK Biobank data, including *SLC41A2*, *STAT1*, *ADAM12*, *KRT5*, and *SH3D21*, also showed moderate to large effect sizes (OR > 2) for HNC risk (Supplementary Fig. S3). Interestingly, predicted damaging missense variants in *SLC41A2, RFX5*, *KLF4*, *XRCC6*, *LEMD3*, and *AK2* exhibited large, statistically significant odds ratios (Supplementary Fig. S5).

**Figure 2.**
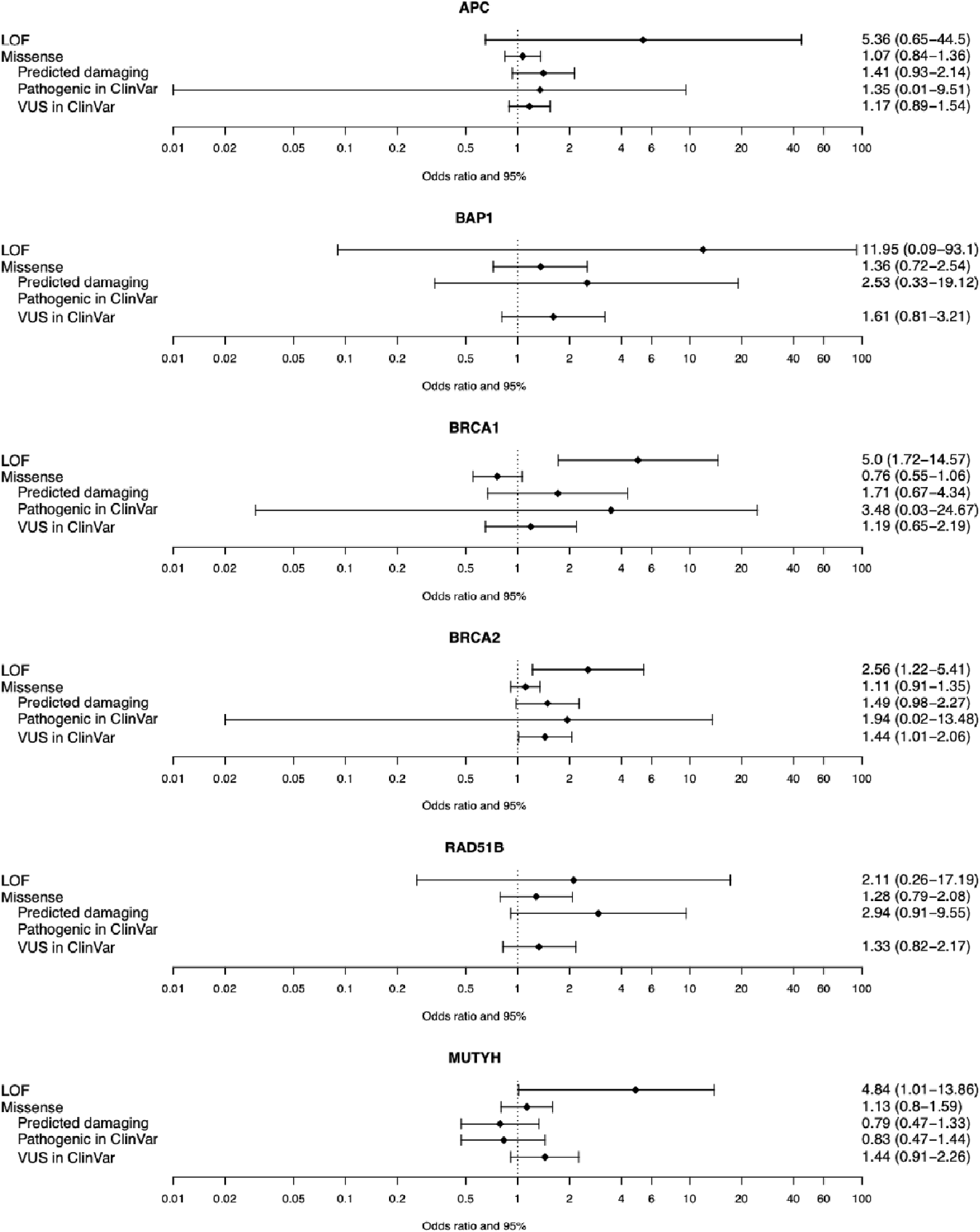
Forest plot of OR of various types of variants in known cancer predisposition genes (with evidence of association of cancers) after combining results from the 3-center data (Houston, North Carolina, and Taiwan) and UK Biobank.

In addition to gene-based analyses, we performed gene set analysis (GSA) to assess the joint effects of biologically correlated genes. We conducted a pathway-based analysis using Fisher’s method to combine the gene-based meta p-values in selected pathways. Three gene sets enriched for DNA homologous recombination repair genes were nominally significant: 1) DNA homologous recombination repair core cancer susceptibility genes, 2) Fanconi anemia complementation gene group, and 3) the homologous recombination repair of replication independent double strand breaks from REACTOME (Table 5). We observed no evidence of association from the following Gene Ontology (GO) sets of a priori interest: GO: negative regulation of endothelial cell migration, GO: stem cell division, and GO: regulation of fibroblast migration, nor from the DNA mismatch repair core cancer susceptibility genes, which served as a negative control.

**Table 5.** Pathway analysis for pathways with p-value <0.25 for the association with head and neck cancer risk.

| Pathway | Genes | Meta p-value |
| --- | --- | --- |
| DNA homologous recombination repair core susceptibility genes | *BRCA1,BARD1,BRCA2,PALB2,RAD51B,RAD51C* | 0.0004 |
| HGNC 548: Fanconi anemia complementation groups | *BRCA1,BRCA2,BRIP1,ERCC4,FANCA,FANCB,FANCC,FANCD2,FANCE,FANCF,FANCG,FANCI,FANCL,FANCM,MAD2L2,PALB2,RAD51,RAD51C,SLX4,UBE2T,XRCC2,RFWD3* | 0.0364 |
| REACTOME: homologous recombination repair of replication independent double strand breaks | *RAD50,H2AX,LIG1,MRE11,NBN,ATM,RAD51,RAD52,RPA1,RPA2,RPA3,BRCA1,BRCA2,TP53BP1,BRIP1,MDC1* | 0.0431 |
| GO: negative regulation of endothelial cell migration | *THBS1,KLF4,DAB2IP,APOH,TGFB1,SP100,SLIT2,GDF2,ITGB1BP1,ANGPT2,CXCL13,STARD13,VASH1,ACVRL1,DLL4,PTPRM,PDCD10,AGTR2,RGCC,CSNK2B,DCN,ANGPT4,HMGB1,APOE,FGF2,SYNJ2BP,SERPINF1,STC1,MAP2K5,HDAC5,RHOA,SVBP,NOTCH1,NR2F2,HRG,BMP10,AGER,KRIT1,MMRN2* | 0.0691 |
| GO: stem cell division | *PAFAH1B1,FZD7,ZFP36L2,CDKN2A,WNT3A,TEAD3,NUMBL,TIAL1,TGFB2,FGF13,LEF1,WNT7A,DCT,NUMB,FGFR2,KIT,ASPM,WWTR1,VANGL2,ETV5,NOTCH1,HOXB4,ING2,CUL3,DOCK7,RAB10,FGFR1,SOX5,ZBTB16* | 0.1552 |
| GO: regulation of fibroblast migration | *BAG4,UTS2,PRR5L,RCC2,RAC1,GNA12,SDC4,AGER,DMTN,FER,WDPCP,DDR2,SLC8A1,HAS1,BRAF,TGFB1,RFFL,CYGB,ARHGAP4,THBS1,AKT1,ARHGEF7,HYAL2,PRKCE,FGF2,MTA2,ITGB1BP1,CORO1C* | 0.2289 |
| Negative control: DNA mismatch repair core susceptibility genes | *MLH1,MSH2,MSH6,PMS2* | 0.9732 |

We further generated pathway-based rare variant effect size estimates, combining LoF and predicted damaging variants for HNC risk. Separate analyses by population were followed by a meta-analysis which further supported the effect size of these pathways. The DNA homologous recombination repair pathway had an OR of 1.67 (95% CI: 1.26-2.22) with both LoF and predicted damaging variants and an OR of 1.60 (95% CI: 0.89-2.79) with LoF variants alone. We observed no evidence of association in other pathways including negative regulation of endothelial cell migration, stem cell division, and regulation of fibroblast migration. (Supplementary Tables S2 to S4)

We used the Synergy Index to investigate whether there were interactions between the HNC susceptibility genes with cigarette smoking. *APC, BRCA2*, and the DNA repair gene set exhibited borderline interactions with smoking, with synergy indices and 95% confidence intervals of 1.90 (95% CI: 0.98-3.70), 1.75 (95% CI: 0.90-3.42), and 1.56 (95% CI: 0.97-2.53), respectively (Table 6 and Supplementary Table S5).

**Table 6.** Test for interaction on the additive scale between cigarette smoking and pathway and genes on the risk of head and neck cancer using LOF and predicted damaging variants, combining results from the 3-center data (Houston, North Carolina, and Taiwan) and UK Biobank*.

| Gene/pathway | Variant Type | OR^ for smoking | OR for genes | OR for both | Synergy Index | One-sided p-value |
| --- | --- | --- | --- | --- | --- | --- |
| *DNA homologous recombination repair core susceptibility genes* | LoF only | 2.95[2.64,3.30] | 2.04[0.82,4.63] | 6.25[2.99,12.2] | 1.75[0.64,4.80] | 0.137 |
|  | LoF and PDM** | 2.97[2.65,3.33] | 1.74[1.14,2.62] | 5.24[3.74,7.32] | 1.56[0.97,2.53] | 0.034 |
| *APC* | LoF only | 2.95[2.64,3.29] | 47.2[0.36,418.] | 22.1[0.36,207.] | 0.44[0.01,26.08] | 0.654 |
|  | LoF and PDM | 2.92[2.62,3.27] | 1.13[0.53,2.19] | 4.90[3.14,7.50] | 1.90[0.98,3.70] | 0.029 |
| *BAP1* | LoF only | 2.95[2.64,3.30] | 23.2[0.18,188.] | 82.8[0.60,1040.] | 3.39[0.05,209.59] | 0.281 |
|  | LoF and PDM | 2.95[2.64,3.30] | 1.72[0.28,8.98] | 6.78[1.35,26.3] | 2.17[0.30,15.81] | 0.222 |
| *BARD1* | LoF only | 2.96[2.65,3.30] | 3.11[0.43,20.6] | 2.41[0.49,12.0] | 0.35[0.02,6.45] | 0.761 |
|  | LoF and PDM | 2.97[2.66,3.32] | 3.01[0.94,8.80] | 3.51[1.14,10.2] | 0.63[0.12,3.45] | 0.702 |
| *BRCA1* | LoF only | 2.96[2.65,3.31] | 14.3[1.82,52.3] | 7.35[1.04,36.5] | 0.42[0.04,4.84] | 0.758 |
|  | LoF and PDM | 2.95[2.64,3.30] | 1.70[0.55,4.50] | 5.09[2.53,10.0] | 1.55[0.53,4.51] | 0.213 |
| *BRCA2* | LoF only | 2.93[2.63,3.28] | 0.87[0.16,3.23] | 10.8[3.63,26.1] | 5.41[1.52,19.26] | 0.005 |
|  | LoF and PDM | 2.93[2.62,3.28] | 1.16[0.62,2.07] | 4.67[2.95,7.42] | 1.75[0.90,3.42] | 0.049 |
| *MUTYH* | LoF only | 2.93[2.63,3.28] | 0.38[0.040,1.73] | 4.26[1.74,11.2] | 2.48[0.68,9.12] | 0.085 |
|  | LoF and PDM | 2.93[2.63,3.28] | 0.54[0.17,1.38] | 2.64[1.60,4.32] | 1.11[0.46,2.67] | 0.408 |
| *PALB2* | LoF only | 2.94[2.64,3.29] | 0.56[0.0039,6.87] | 4.20[0.56,18.9] | 2.12[0.18,25.30] | 0.276 |
|  | LoF and PDM | 2.95[2.64,3.30] | 1.99[0.44,7.37] | 7.74[2.07,22.0] | 2.30[0.46,11.41] | 0.155 |
| *RAD51B* | LoF only | 2.95[2.64,3.29] | 4.71[0.037,33.0] | 8.05[0.33,59.8] | 1.25[0.03,45.19] | 0.452 |
|  | LoF and PDM | 2.95[2.64,3.30] | 2.42[0.51,8.96] | 8.32[2.74,23.0] | 2.17[0.47,10.15] | 0.162 |
| *RAD51C* | LoF only | 2.96[2.65,3.31] | 30.5[3.53,118.] | 46.6[5.05,194.] | 1.45[0.14,14.76] | 0.378 |
|  | LoF and PDM | 2.97[2.65,3.31] | 5.36[1.33,20.2] | 7.10[1.40,27.6] | 0.96[0.13,7.21] | 0.514 |

^ Showing point estimate and 95% confidence interval
\* Adjusted for race and sex
\*\* PDM: predicted damaging missense

## Discussion

We identified multiple DNA homologous repair pathway genes where LoF mutations had large, statistically significant effect sizes, including *BRCA1*, *BRCA2*, and *RAD51B*. In addition, we identified associations in three additional cancer predisposition genes, *APC*, *BAP1*, and *MUTYH*. *MUTYH* is a recessive cancer predisposition gene leading to colonic adenomatous polyposis and cancer, but there is some speculation that a single altered copy slightly increases risk.^19^ *MUTYH* is involved in oxidative damage repair, which is associated with alcohol and smoking exposures.

Smoking and/or alcohol exposure can cause DNA hypermethylation, DNA double-strand breaks, and DNA oxidative damage, leading to altered expression and/or function of cancer hallmark genes.^20–22^ Inactivation of *APC* gene expression through hypermethylation is a well-established mechanism of oral cancer tumorigenesis.^23^ Consequently, it is understandable that cigarette smokers with an inherited heterozygous pathogenic variant in *APC* might have a higher risk of oral cancers than non-smokers because smoking-associated DNA methylation would only need to inactivate the remaining functional *APC* allele. Based on our results, the LoF variants in *APC* confer an approximate 5-fold increased risk of developing HNC (Figure 2), and there is evidence of an interaction with smoking (Table 6), in accord with the exposure-driven hypermethylation hypothesis.

Similarly, since proteins encoded by the core DNA homologous recombination repair cancer susceptibility genes (*BRCA1*, *BARD1*, *BRCA2*, *PALB2*, and some of the *RAD51* paralogs) play key roles in scar-less repair of DNA double strand breaks, an inherited heterozygous pathogenic variant in any one of these genes could interact with cigarette smoking, resulting in an increased risk of tumorigenesis in directly exposed tissues. We observed that potentially pathogenic variants in an HRR gene confer an approximately 1.64-fold increased risk of HNC (Supplementary Table S2), and there is again evidence of an interaction with smoking (Table 6).

Tobacco smoke contains reactive oxygen species that can cause oxidation of a wide variety of biomolecules, including DNA, RNA, and free nucleotides. Prominent among these oxidation products is 8-oxoguanine (OG), which, if unrepaired, leads to G to A transversions.^24^ *MUTYH* encodes an enzyme involved in base excision repair, removing adenine at inappropriate OG:A base pair.^25^ *MUTYH* is well recognized as a high-risk recessive dominant susceptibility gene for polyposis colorectal cancer^26^ and modest-risk dominant CRC susceptibility gene.^27,28^ We observed borderline evidence that heterozygous LoF variants in *MUTYH* are involved in an Ottman Model B gene-environment interaction^29^ (P = 0.085) wherein the increased risk conferred by LOF variants seems to be modulated entirely by smoking status, with LOF variants not conferring an increase in risk to non-smokers (OR=0.38, 95% CI: 0.04-1.73), but with a substantial increase in risk among smokers (smoker and LOF variant carrier OR = 4.26, 95% CI: 1.74-11.2 versus OR = 2.93, 95% CI: 2.63-3.28 among smokers without a LOF variant).

A major strength of this study was the large heterogeneous cohort through collaboration across multiple sites in the US and the international collaboration with Taiwan. Additional strengths include the familial design in Phase I, which resulted in a small panel of 501 genes enriched for HNC susceptibility genes. Across Phase 2 and the UK Biobank, we employed a rigorous case-control study design, matching cases and controls by sex, age, race, ethnicity, recruitment center, as well as sequencing center, platform, and protocol. By combining tests that weight variants by *in silico* estimates of functional severity together with unweighted approaches that assign equal weight to all variants, our gene-based approach was sensitive to a broad variety of genetic architectures.

One limitation of this study was the lack of HPV data, which is recognized an important risk factor for oropharyngeal cancer. We were able to mitigate this limitation through oropharyngeal cancer subsite analysis, though our sample sizes and statistical power were limited (Supplementary Tables S6 to S9). Although our overall sample size included approximately 3,600 HNC cases, sample availability varied widely across populations, resulting in limited statistical power to detect population-specific associations. Additional studies are needed to increase sample sizes for specific cancer subsites and across multiple populations.

## Conclusion

We observed associations with head and neck cancer risk for six known cancer susceptibility genes (*BRCA1*, *BRCA2*, *RAD51B*, *BAP1*, *APC*, and *MUTYH*), in addition to an overlapping set of six core DNA homologous recombination repair pathway cancer susceptibility genes (*BRCA1*, *BARD1*, *BRCA2*, *PALB2*, *RAD51B*, and *RAD51C*) that are most closely identified with hereditary breast and ovarian cancer. We also identified potential interactions between these genes and cigarette smoking. We estimate that smokers with pathogenic variants in either *APC* or one of the DNA homologous recombination repair genes are at a 4- to 5-fold increased risk of developing head and neck cancer. Further studies are needed to confirm these associations and to generate more precise risk estimates.

**Figure S1.**
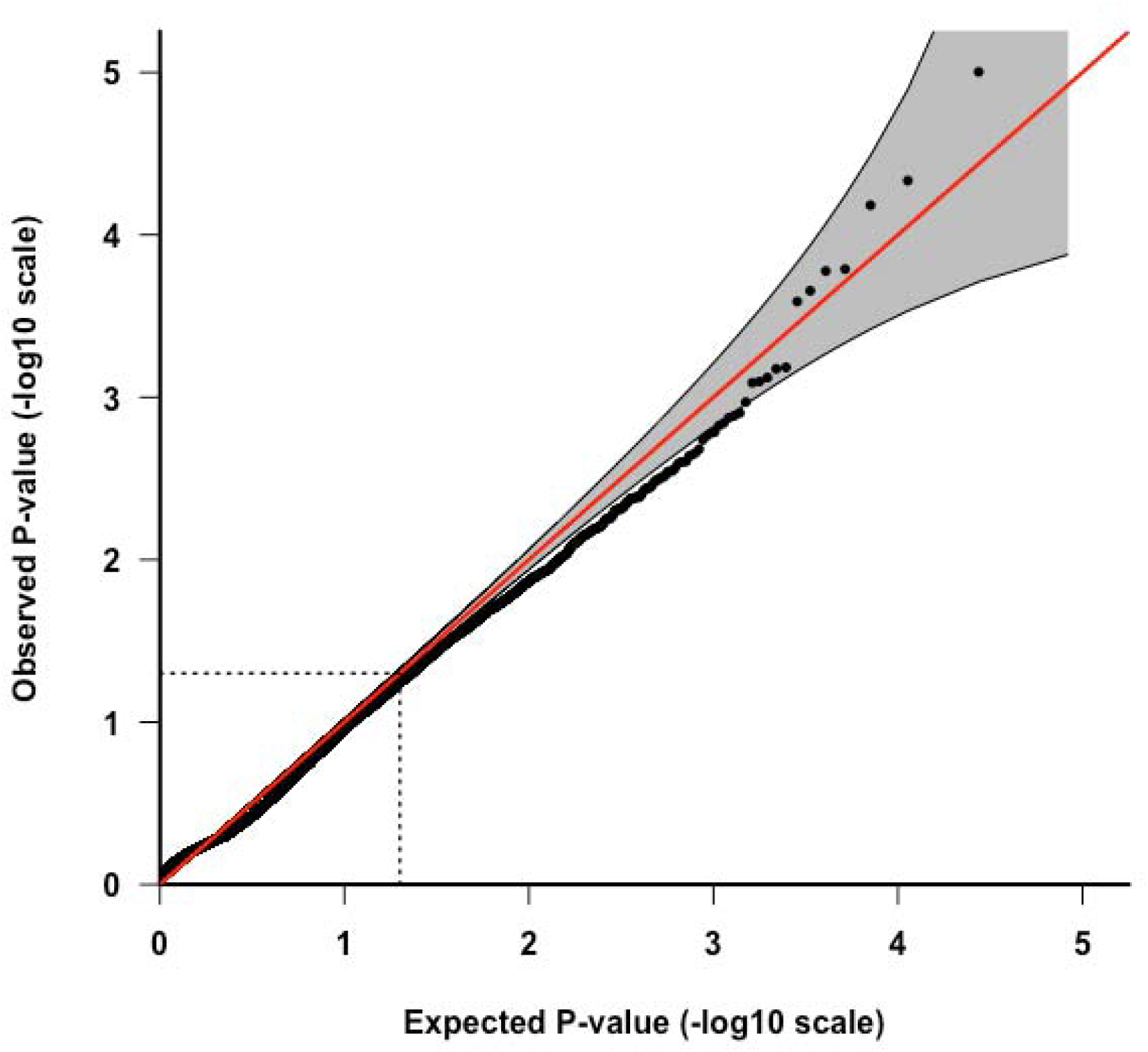
Q–Q plot of Meta P-value for gene-based association analysis for head and neck cancer risk.

**Figure S2.**
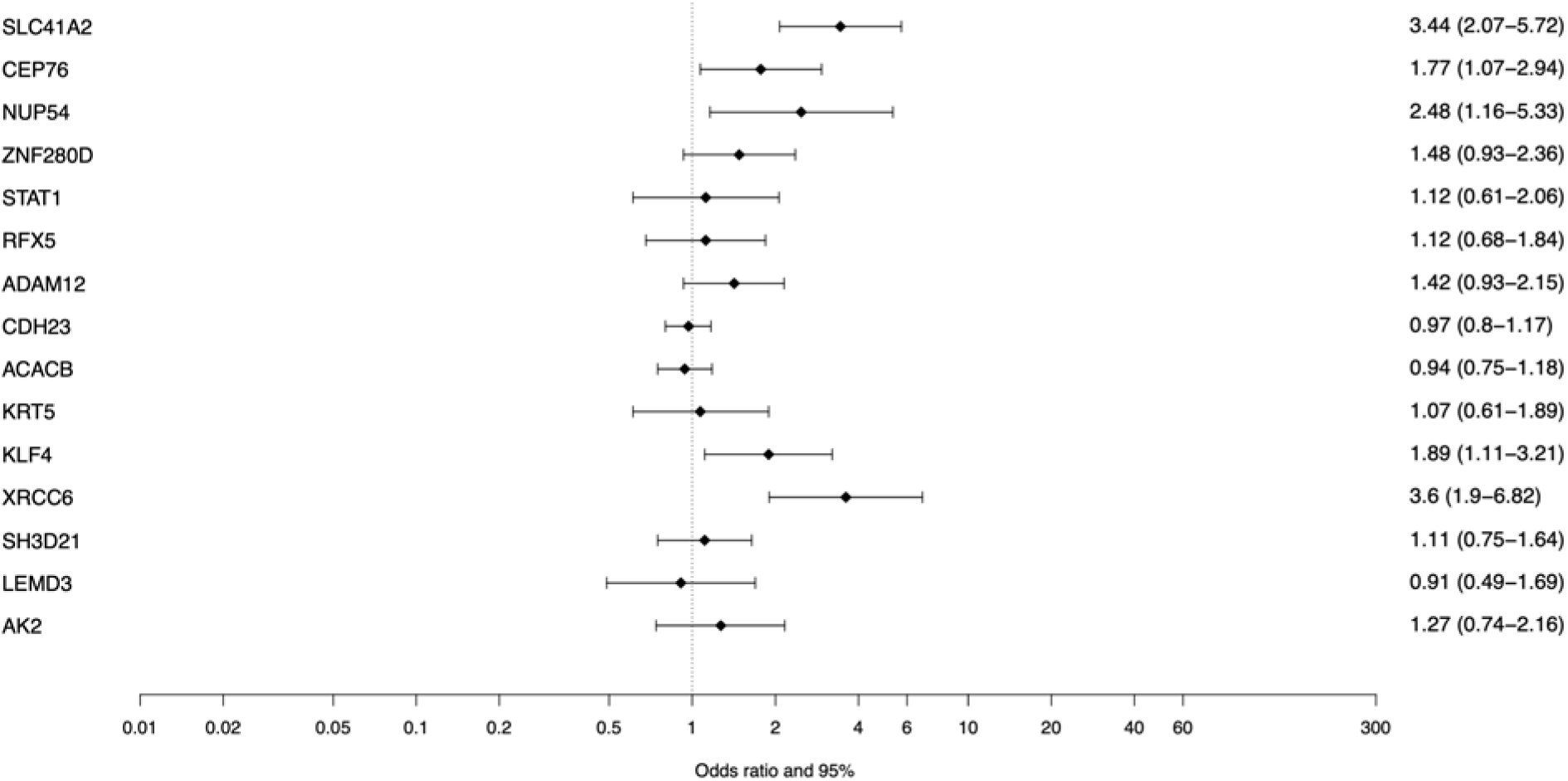
ORs of all variants in for genes with Meta p-value < 0.05 after combining results from Houston, North Carolina, Taiwan, and UK Biobank in non-Hispanic European population.

**Figure S3.**
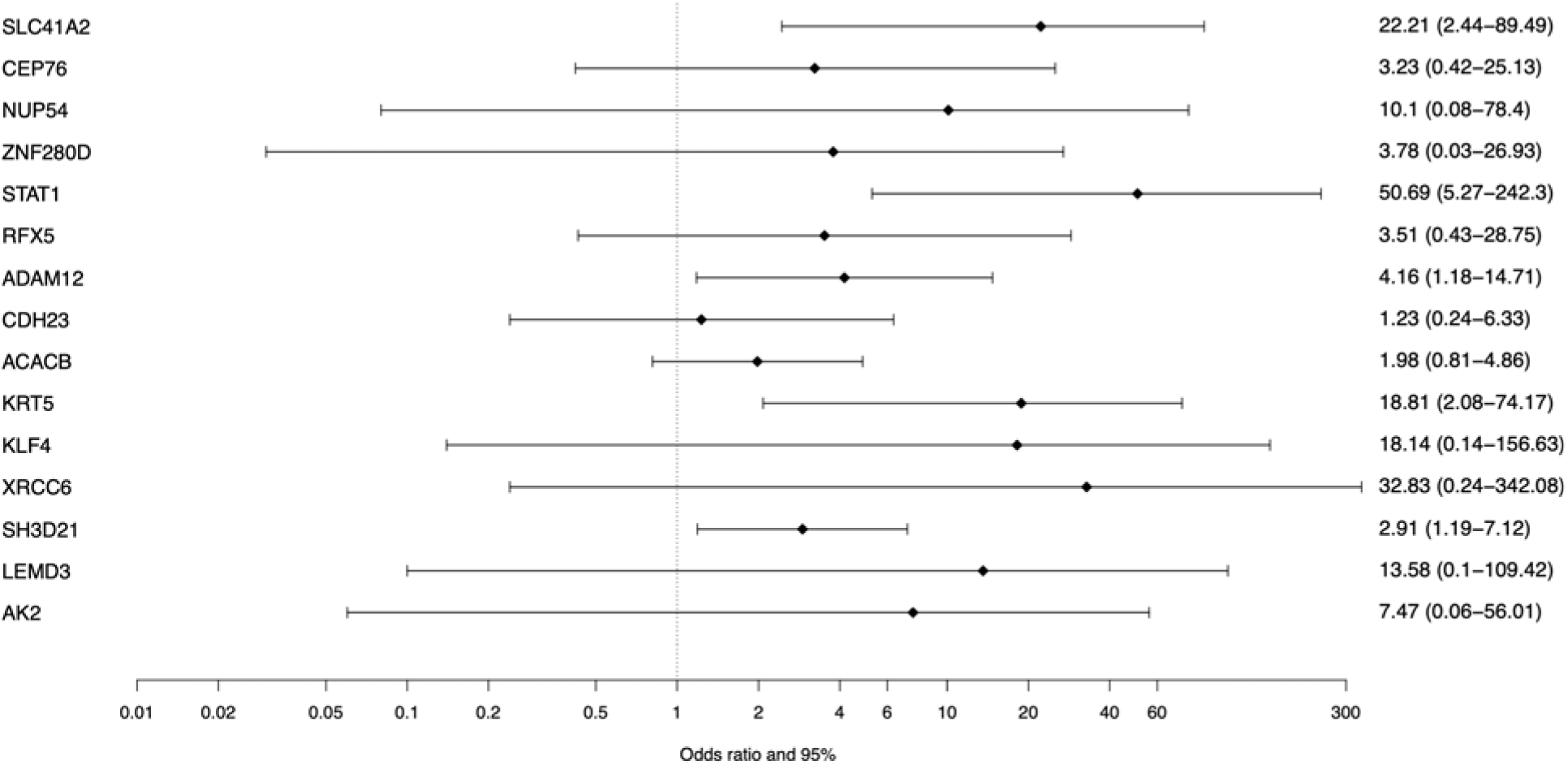
ORs of LOF variants in for genes with Meta p-value < 0.05 after combining results from Houston, North Carolina, Taiwan, and UK Biobank in non-Hispanic European population.

**Figure S4.**
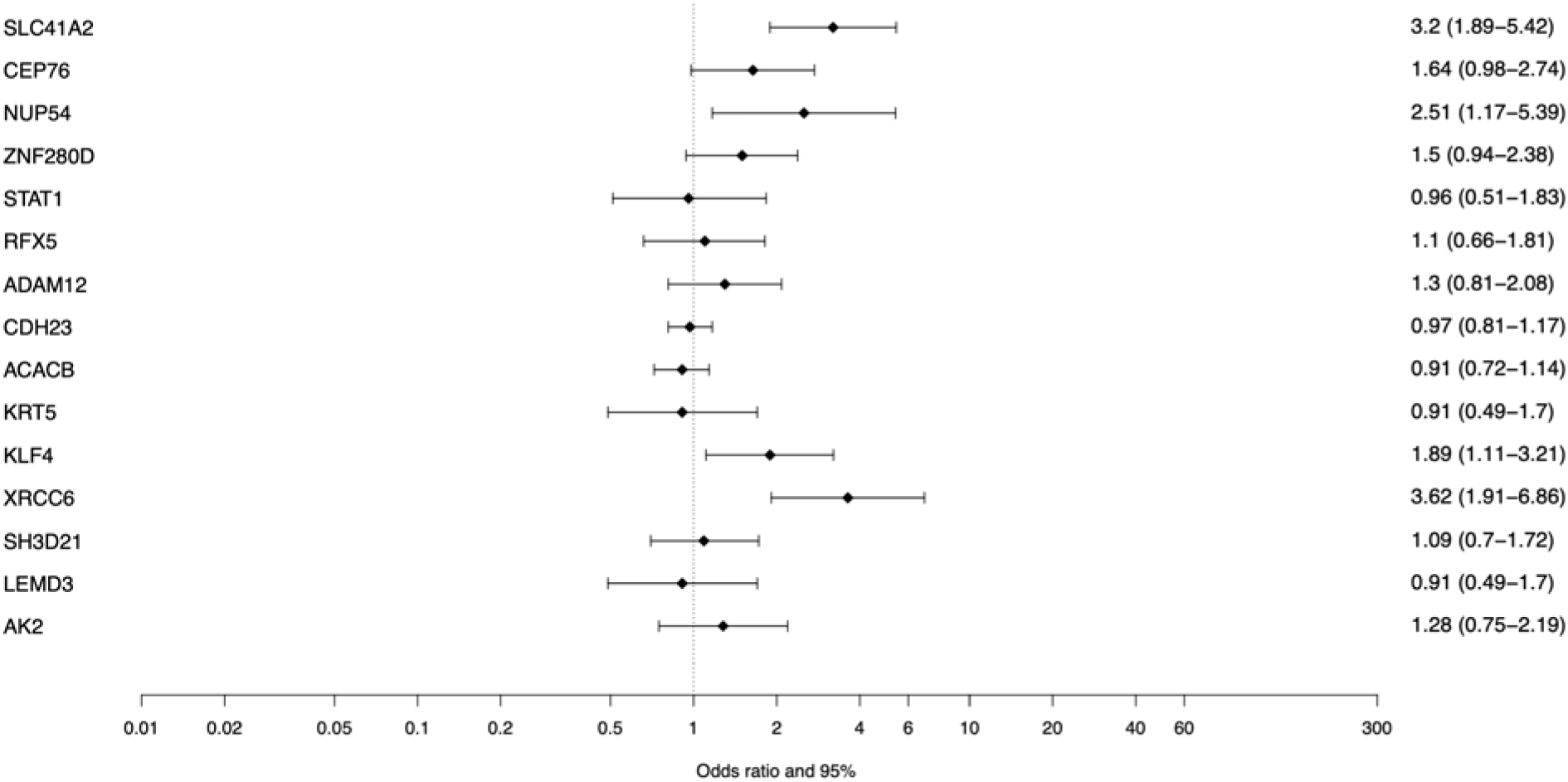
ORs of all missense variants in for genes with Meta p-value < 0.05 after combining results from Houston, North Carolina, Taiwan, and UK Biobank in non-Hispanic European population.

**Figure S5.**
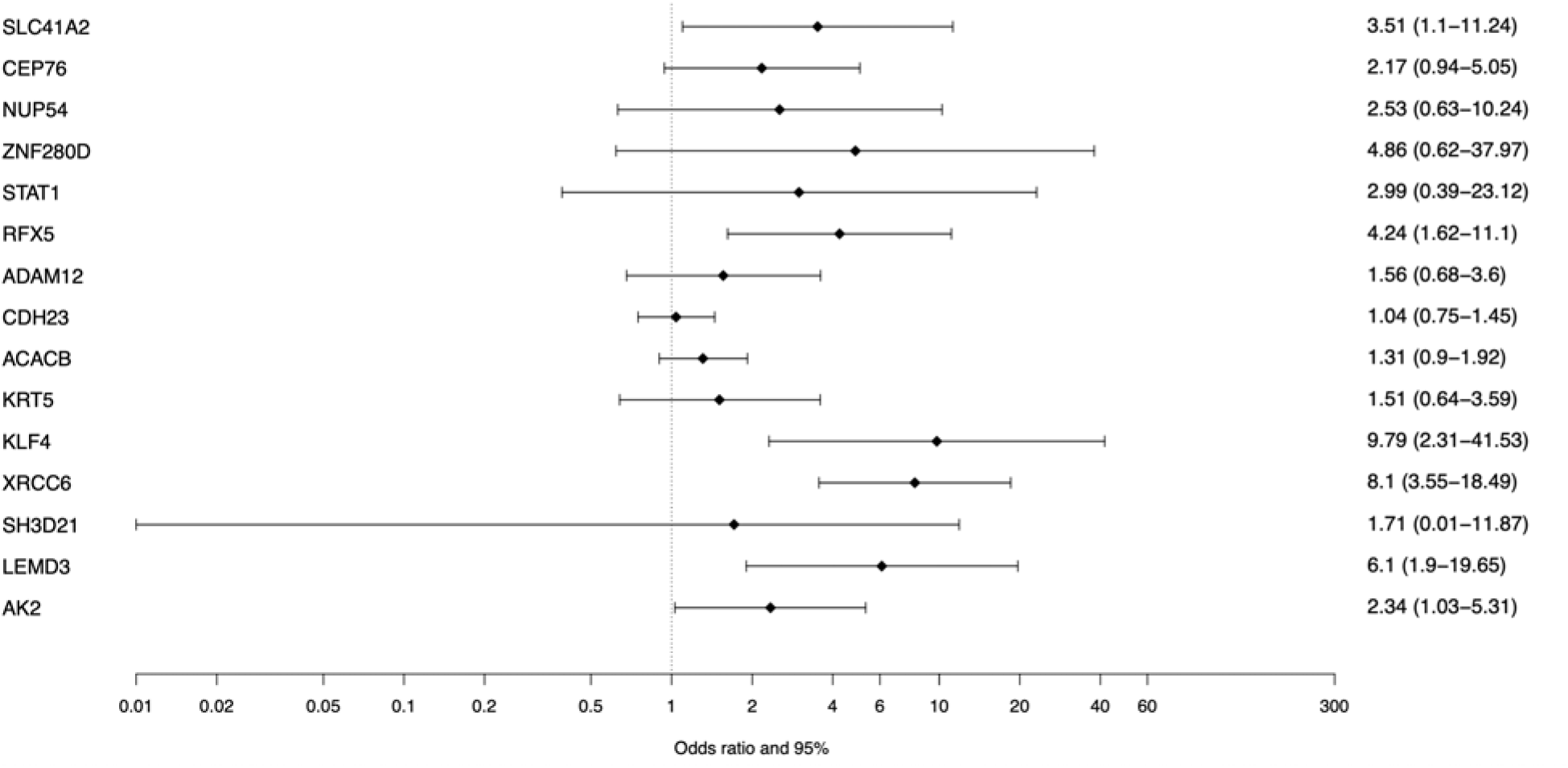
ORs of predicted pathogenic missense variants in for genes with Meta p-value < 0.05 after combining results from Houston, North Carolina, Taiwan, and UK Biobank in non-Hispanic European population.

## Material and Methods

### Study cohorts of head and neck cancer

In this study, we conducted a multi-center case-control analysis for HNC. HNC cancer subsites were defined by the International Classification of Diseases for Oncology, Third Edition (ICD-O-3) codes. Codes for oral cavity were C00.3, C00.4, C00.5, C00.6, C00.8, C00.9, C02.0, C02.1, C02.2, C02.3, C03.0, C03.1, C03.9, C04.0, C04.1, C04.8, C04.9 C05.0, C06.0, C06.1, C06.2, C06.8, and C06.9. Codes for oropharynx were C01.9, C02.4, C05.1, C05.2, C09.0, C09.1, C09.8, C09.9, C10.0, C10.2, C10.3, C10.4, C10.8, and C10.9. Codes for hypopharynx were C12.9, C13.0, C13.1, C13.2, C13.8, and C13.9. Codes for larynx were C10.1, C32.0, C32.1, C32.2, C32.3, C32.8, C32.9. Codes for oral cavity-oropharynx-hypopharynx, not otherwise specified (NOS) were C02.8, C02.9, C05.8, C05.9, C14.0, C14.2, C14.8.

The sample recruitment of this study consisted of two phases. In Phase 1, we included only familial HNC cases from study centers in Houston, North Carolina, Taiwan, and Utah. Familial HNC cases were defined as patients who had first- or second-degree family members diagnosed with HNC. In Phase 2, we included non-familial HNC cases and controls from the study centers in Houston, North Carolina, and Taiwan. The North Carolina study was a population-based case-control study with recruitment from 2002-2006.^30^ The Houston study was a hospital-based case-control study with recruitment from 2001-2006 based at the MD Anderson Cancer Center.^31^ The Taiwan HNC patients were from a multicenter study conducted from 2010-2015.^32^ The patients in Utah were identified through the Utah Population Database.^33^ Demographic characteristics and lifestyle factors were collected via questionnaires administered by trained interviewers at each study center. DNA was extracted from blood samples. In Phase 1, the final analysis included 220 familial HNC cases. In Phase 2, there were 2,134 HNC cases and 2,072 controls. The study was approved by Institutional Review Boards at each of the institutes involved. To perform a meta-analysis, we obtained whole exome sequencing data from UK Biobank (200K release), which included 531 HNC patients per the selection criteria above and 119,716 cancer-free controls.

### Whole exome sequencing of Phase 1 samples

We first conducted whole exome sequencing (WES) using Phase 1 samples with an average depth of over 60x using Illumina HiSeq 2500 at the HCI High-Throughput Genomics Core Facility at the University of Utah Health Sciences. The FASTQ reads were mapped to the human genome reference GRCh37 downloaded from the GATK resource using BWA-MEM (v0.7.10).^34^ Duplicates were marked using Samblaster (v0.1.22)^35^, sorted, and formatted with Sambamba^36^. We also obtained WES data of controls from the 1000 Genomes Project.^10^ The 1000 Genomes samples included non-Finnish Europeans (NFE), African (AFR), and East Asian (EAS) descendants to match with the cases. We conducted individual variant calling using GATK HaplotypeCaller and used CombineGVCFs to create genome VCF files, followed by joint genotype using Sentieon© GVCFTyper^37^ calling for all samples, and recalibrated the quality of variant calls following GATK’s best practice.^16^

We then conducted variant and sample level quality controls by using the following criteria: (1) Exclude genotypes with read depth <10x or genotype quality <55. (2) Exclude variants that failed in Variant Quality Score Recalibration (VQSR), missing rate >2%, less than 80% of samples with depth <10x, the mean sequencing depth was <10 or >500, or the proportion of heterozygous genotypes among non-missing haploidy individuals >10%. (3) Exclude samples with missing rate >5%, Het/Hom ratio >5, Ti/Tv ratio <2.5, the mean read depth <20x, the mean genotype quality is <55, or conflicted genetic and self-reported sex. Cryptic relatedness among samples were identified by the program KING. The individuals with the lowest missing rate among each group of duplicated or related samples were retained for subsequent analysis. We performed principal component analysis (PCA) to control for population stratification. We assessed whether the cases and controls were comparable by testing the differences in the number of rare variants per sample.

We annotated BayesDel score as a measure of the deleteriousness of each variant in the association test. We annotated functional consequences of variants against the RefSeq database, restricting to principal transcripts obtained from the APPRIS (annotation of principal and alternative splice isoforms) database. LoF variants included FrameShifting, StopGain, and SplitSite variants. Any variant located in the last 5% of a coding region were not considered as LoF. Variants located within two base pairs in an intron were considered as SplitSite variants. For variants at -3 to +8 base pairs around a splicing donor site or at -12 to +2 base pairs around a splicing acceptor site, the splicing alternating effect was obtained from the dbscSNV database with the cutoff value of 0.9. We also conducted clinical annotation using the ClinVar database and excluded variants classified as benign. We conducted a gene-based burden test using a one-sided Fisher’s Exact test within each population, restricting to rare variants with minor allele frequency or MAF <=0.01), and then a combined p-value was calculated by Fisher’s method weighted by sample size. Additionally, we conducted a gene prioritization analysis combining gene-based burden test p-values with biological relevant scores.

To extend the sample size, we further conducted a case-control analysis including additional HNC samples from the Cancer Genome Atlas Head-Neck Squamous Cell Carcinoma (TCGA-HNSC) data, cancer-free controls recruited and sequenced at University of Texas MD Anderson Cancer Center, and unaffected parents from the National Database for Autism Research (NDAR). We conducted reference genome alignment and jointly genotype calling following GATK’s best practice. This dataset contains data from multiple studies with heterogeneity in target capture and sequencing technologies, often introducing strong technological stratification biases that overwhelm subtle signals of association tests. Thus, we conducted sample level and variant level quality control (QC) using the Cross-Platform Association Toolkit (XPAT) with default parameters. We grouped 731 TCGA cases and 4988 NDAR controls according to their predicted population in PCA (including non-Hispanic European, African American, East Asian, and Latin/admixed American), followed by a gene-based association analysis using VAAST. We conducted gene-based association test for each population group and combined the summary statistics in a meta-analysis weighted by sample size. After that, we used PheVor to combine biomedical ontologies to identify HNC-associated genes.

Next, we developed a targeted sequencing panel for HNC using Phase 1 data. As described above, we conducted multiple analyses: (1) gene-based association test; (2) gene-set-based association test; and (3) gene prioritization combining gene-based burden test results and biological relevant scores. Each analysis was conducted on two sets of data where the control samples were different. We then selected the top ∼100 genes from each analysis, resulting in 417 unique genes. Subsequently, we hand-picked 84 additional cancer-related genes to make a 501-gene panel for the Phase 2 targeted sequencing.

### Targeted gene sequencing of Phase 2 samples

Utilizing the panel of HNC risk genes designed using Phase 1 data, we performed targeted gene sequencing for all Phase 2 samples recruited from the three centers. The experiment was also conducted by the HCI High-Throughput Genomics Core Facility at the University of Utah Health Sciences. After obtaining the sequencing reads, we first trimmed FASTQ files using Cutadapt^38^ to remove residual adapters in the tails. We used FLASH^39^ to combine the overlapped tail of the paired-end reads into a single long read if possible. Uncombined reads were treated as two single-end reads in the following analysis. The GATK Best Practices workflow^16^ was applied for reference genome alignment, joint variant genotype calling, and variant quality score recalibration. The sequencing reads were mapped to the human reference genome (University of California, Santa Cruz or UCSC HG19) using Burrows-Wheeler Aligner (BWA) (v0.7.9a).^34^ We removed unmapped reads, reads with mapping quality equal to 0, and duplicated reads using Picard tools (http://broadinstitute.github.io/picard). We conducted local realignment and minimized mismatched bases across reads using GATK’s IndelRealigner. We recalibrated the base quality score and generated the final clean BAM file using GATK’s BaseRecalibrator. We conducted joint variant genotype calling of all samples using GATK’s HaplotypeCaller and variant quality score recalibration (VQSR) using GATK’s VariantRecalibrator. To ensure the quality of the genotypes, we compared the results from the above two pipelines and only included the consistent genotype in the subsequent analysis. For UK BioBank samples, we downloaded the individual gVCF files generated from whole exome sequencing (WES) from UK BioBank database. We utilized DeepVariant, which was implemented in GLnexus, to perform joint genotype calling.^18^

After joint genotype calling, we conducted sample level and variant level quality control (QC) using the Cross-Platform Association Toolkit (XPAT) for each dataset separately.^13^

For sample-level quality control (QC), we computed the ratio of heterozygous variant counts to homozygous variant counts on autosomal chromosomes for each sample. Samples with ratios exceeding 3 were excluded. Additionally, we calculated NC90 scores for each sample using XPAT. NC90 scores represent the proportion of missing genotypes in a sample among all variants with call rates of 90% or greater across all samples, limited to variants within the target regions. Samples with NC90 scores greater than 0.05 were excluded. We also removed samples identified as 1st and 2nd-degree relatives predicted by RELPAIR.^40^ Furthermore, we conducted Principal Component Analysis (PCA) to project all cases and controls onto a reference panel constructed using 1000 Genomes Phase 3 data.^10^ This step was performed to identify genetically matched cases and controls for the subsequent association tests. For the target sequencing data of samples from the three centers, we grouped the samples into four ethnicity groups: non-Hispanic European, African American, Asian, and Hispanic. For UK Biobank data, we only selected cases and controls of non-Hispanic European ancestry.

We performed variant-level quality control (QC) for each population group of the three center samples and UK Biobank data, using XPAT with default parameters unless otherwise specified. Variants with VQSR tranche sensitivity scores >99.95 for SNPs and >99.5 for INDELs were excluded. Additionally, variants with an average allele fraction lower than 20% in our data were excluded. The average allele fraction was calculated as the proportion of reads supporting the minor allele among all reads in each sample with the minor allele. Variants with a missing genotype rate exceeding 10% across all samples were also excluded. Furthermore, variants with a p < 10^-6^ in a Hardy-Weinberg equilibrium test in controls were excluded from further analysis. We conducted an internal PCA using cases and controls of each population and dataset separately, using well-behavior markers from the corresponding population (MAF > 20%), to capture population stratification within each group. The first 10 components will be used as covariant in the following subsequent association tests.

For UK BioBank samples, we downloaded the individual gVCF files generated from whole exome sequencing (WES) from UK BioBank database. We utilized DeepVariant, which was implemented in GLnexus, to perform joint genotype calling.12 After joint genotype calling, we conducted sample level and variant level quality control (QC) using XPAT with the same parameters above.19 We further removed samples identified as close relatives predicted by UK Biobank. We used the first 10 components for UK Biobank’s PCA analysis as covariates in the subsequent association tests.

### Gene-based association analysis

For each of the four populations of the three center samples and the non-Hispanic European group of UK Biobank samples, we conducted the gene-based association test using three approaches. (1) We first conducted gene-based association tests with all rare variants (minor allele frequency or MAF < 0.005) using the Variant Annotation, Analysis and Search Tool (VAAST 2).^14^ The analysis considered all coding variants, including missense, stop gains, stop losses, splicing variants, INDELs, and CNVs, weighting each variant according to the estimated degree of protein dysfunction conferred based on the CASM score in VAAST. We only tested genes with at least four scorable variants. Among the 501 genes included in Phase 2 targeted sequencing panel, 461 genes were tested using the 3-center data. (2) Next, we performed gene-based association analyses using logistic regression, first with LoF variants only. (3) We then performed gene-based association analyses using logistic regression with a combination of LoF and predicted damaging variants. As described above, we used BayesDel score to estimate the deleteriousness of each variant. For all gene-based tests, we used the first ten PCs, age, and sex as covariates to control for population and technological stratification. For each population group, we conducted an aggregated Cauchy association test (ACAT)^41^ to combine the p values from individual tests. We next conducted a meta-analysis to combine the ACAT p values of four population groups for each gene, using Stouffer’s method. We used efficient sample size from each group as weight in the meta-analysis, defined as 4*number of cases*number of controls divided by the sum of cases and controls. We also conducted pathway-based analysis by combining p values of genes in each candidate pathway using Fisher’s method.

### Effect size estimation for genes and pathways

We evaluated variant effect sizes of top genes in association tests for head and neck cancer risk according to a variety of functional classifications, considering their amino acid changes, *in silico* functional predictions, and variant annotation status in ClinVar using ANNOVAR (Version 20221231).^42^ For ClinVar annotations, we included ‘pathogenic’ and ‘likely pathogenic’ variants in the known pathogenic category, and uncertain or unknown significance as VUS. The effect size was estimated using the Firth regression method for each population group. We used the first ten PCs, age, and sex as covariates. To avoid bias due to the small sample size, we required at least 3 carriers of variants in cases and controls to estimate the odds ratio. For the non-Hispanic European group, we conducted the meta-analysis of the odds ratio to combine the estimates from the 3-center data and UK Biobank data.^43^

### Interaction analysis (Gene x Smoking, Gene x Drinking)

To assess epistasis between two predictors, we coded the two predictors into a four-level variable, corresponding to (1) negative for both predictors, (2) positive for the first predictor, (3) positive for the second predictor, and (4) positive for both predictors, respectively. Logistic regression was performed on this variable, using level 1 as the reference, adjusting for ancestry and sex. Additive interaction between the two predictors was then assessed through the Synergy Index.

## Data Availability

All data produced in the present study are available upon reasonable request to the authors

## Acknowledgements

Research reported in this publication was supported by the NIDCR of the NIH under award number R01DE023414 (Hashibe/Tavtigian). We thank the UPDB staff of Huntsman Cancer Institute, University of Utah (funded in part by the Huntsman Cancer Foundation) for their role in the ongoing collection, maintenance and support of the Utah Population Database (UPDB). We also acknowledge partial support for the UPDB through grant P30 CA2014 from the National Cancer Institute, University of Utah and from the University of Utah’s program in Personalized Health and Utah Clinical and Translational Science Institute. The Utah Cancer Registry is funded by the National Cancer Institute’s SEER Program, Contract No. HHSN261201800016I, the US Centers for Disease Control and Prevention’s National Program of Cancer Registries, Cooperative Agreement No. NU58DP007131, with additional support from the University of Utah and Huntsman Cancer Foundation.

